# Hybrid immunity expands the functional humoral footprint of both mRNA and vector-based SARS-CoV-2 vaccines

**DOI:** 10.1101/2022.06.28.22276786

**Authors:** Paulina Kaplonek, Yixiang Deng, Jessica Shih-Lu Lee, Heather J Zar, Dace Zavadska, Marina Johnson, Douglas A. Lauffenburger, David Goldblatt, Galit Alter

**Affiliations:** Ragon Institute of MGH, MIT, and Harvard, Cambridge, MA, USA; Department of Biological Engineering, Massachusetts Institute of Technology, Cambridge, MA 02142, USA; Department of Pediatrics and Child Health, Red Cross War Memorial Children’s Hospital, University of Cape Town, Cape Town, South Africa; SA MRC Unit on Child and Adolescent Health, University of Cape Town, Cape Town, South Africa; Children’s Clinical University Hospital, Riga, Latvia; Great Ormond Street Institute of Child Health Biomedical Research Centre, University College London, London, UK

## Abstract

The COVID-19 pandemic catalyzed a revolution in vaccine development, leading to the testing and approval of several global vaccine platforms that have shown tremendous promise in curbing the pandemic. Yet, despite these successes, waning immunity, and the emergence of variants of concern linked to rising breakthrough infections among vaccinees, have begun to highlight opportunities to improve vaccine platforms and deployment. Real-world vaccine efficacy has highlighted the reduced risk of breakthrough infection and disease among individuals infected and vaccinated, otherwise referred to as hybrid immunity. Hybrid immunity points to the potential for more vigorous or distinct immunity primed by the infection and may confer enhanced protection from COVID-19. Beyond augmented hybrid induced neutralizing antibody and T cell immune responses, here we sought to define whether hybrid immunity may shape the functional humoral immune response to SARS-CoV-2 following Pfizer/BNT162b2 and Moderna mRNA1273 mRNA-based, and ChadOx1/AZ1222 and Ad26.COV2.S vector-based SARS-CoV-2 vaccination. Each vaccine exhibited a unique functional humoral immune profile in the setting of naïve or hybrid immunity. However, hybrid immunity showed a unique augmentation in S2-domain specific functional humoral immunity that was poorly induced in the setting of naïve immune response. These data highlight the immunodominant effect of the S1-domain in the setting of natural immunity, which is highly variable during viral evolution, and the importance of natural infection in breaking this immunodominance in driving immunity to the S2 region of the SARS-CoV-2 S2 domain that is more conserved across variants of concern.

## Introduction

The emergence of SARS-CoV-2 in late 2019 launched an unparalleled global pandemic, causing nearly half a billion documented infections and over 6 million deaths (1). The unpredictable trajectory of disease severity drove the urgent need for vaccine development, which proceeded at an unprecedented speed, leading to the authorization and licensure of several new vaccine platforms (2). Specifically, while both the Pfizer/BioNTech and Moderna mRNA vaccines were associated with greater than 90% protection against severe disease and death (3, 4), the AstraZeneca chimpanzee adenovirus (ChadOx1/AZ1222) was associated with 75% protection (5), and the Johnson & Johnson–Janssen adenovirus 26 (Ad26.COV2.S) was associated with 65% protection (6). Neutralizing antibodies were linked to protective immunity across all platforms during the initial Phase 2b/3 trials when the D614G strain dominated the global pandemic (3-6). However, the emergence of several neutralization resistant variants of concern (VOCs), in the setting of rapidly waning neutralizing antibodies, caused widespread infection in the absence of a proportional increase in severe disease and death. Instead, these data suggest that alternate vaccine-induced immune responses, including T cells (7-10) and non-neutralizing antibody functions (11, 12), likely contribute to vaccine-induced attenuation of disease. However, while T cells are induced variably by these four vaccine platforms, whether non-neutralizing antibodies differ among vaccine platforms and contribute differentially to protection against severe disease and death remains unclear.

Beyond the robust correlation between neutralizing antibody titers and vaccine efficacy in early efficacy trials (13-15), meta-analyses across vaccine trials pointed to a stronger correlation between antibody binding titers and efficacy between vaccine platforms (16, 17). These data pointed to the possibility that additional functions of antibodies, beyond their ability to bind and block infection, may play a critical role in the attenuation of disease. Along these lines, cytotoxic functions of antibodies were linked to survival in a large convalescent plasma therapeutic trial (18), antibody Fc-effector functions are key to the therapeutic activity of several monoclonal therapeutics (19), and the opsonophagocytic function of antibodies are key predictors of survival of severe natural infection (20, 21). However, whether these functions are tuned distinctly across vaccine platforms or linked to the protection afforded by specific vaccines remains unclear.

Real-world efficacy has begun to increase vaccine effectiveness among individuals who were previously infected and then vaccinated, also referred to as hybrid immunity (22, 23). Moreover, several studies have suggested that hybrid immunity is associated with improved protection against multiple VOCs (24-26), including Omicron (27, 28), via induction of antibodies with increased potency and breadth. Additional studies have revealed a more robust spike-specific antibody production (29-31) and more vigorous T cell responses (32, 33) in the setting of hybrid immunity. However, whether hybrid immunity also alters the functional character of the humoral immune response remains unclear.

Preliminary data has pointed to subtle differences in the vaccine-induced antibody profiles between the Pfizer/BioNTech BNT162b2 and Moderna mRNA1273 mRNA vaccines (12). However, whether these functional humoral responses differ among adenovirus viral vaccines as well, and whether they are tuned in the setting of hybrid immunity, has remained unclear. Thus, here we deeply profiled the functional humoral immune response across four novel SARS-CoV-2 vaccine platforms, including the Pfizer/BioNTech BNT162b2 and Moderna mRNA1273 mRNA vaccines, and the AstraZeneca ChadOx1/AZ1222 and Janssen Ad.26COV2.S vaccines. Peak immunogenicity profiles were compared in naïve individuals (without prior SARS-COV-2 infection) and those with previous SARS-CoV-2 infection (hybrid immunity). Striking differences were noted in the functional humoral immune response between mRNA and vectored vaccines in the naïve population that were linked to differences in protective immunity between vaccine platforms. Moreover, we observed both significant increases in the magnitude and quality of the hybrid immune response across the different vaccines, marked by a unique, selectively expanded S2-specific effector antibody response in the setting of hybrid immunity. Given the greater conservation in S2 across variants of concern and across additional coronaviruses, this uniquely expanded S2-specific functional humoral immune response may represent a key mechanism that provides enhanced real-world efficacy against severe disease and death. Thus, next generation vaccines able to promote enhanced functional S2-specific humoral immunity may provide enhanced protection against severe disease and death.

## Results

### Characteristics of the Four Vaccine Cohorts

Serum samples from adults with and without prior SARS-CoV-2 infection who followed a complete immunization schedule using one of the four available SARS-CoV-2 vaccines were analyzed. Patients received either two doses of one of the mRNA vaccines (1) BNT162b2 (Pfizer, vaccination only n = 35, and hybrid immunity n = 9) or (2) mRNA-1273 (Moderna, vaccination only n = 19, and hybrid immunity n = 1); (3) one dose of the Ad26 vectored vaccine Ad26.COV2.S (Janssen, vaccination only n = 25, and hybrid immunity n = 8); or (4) two doses of the ChAdOx vectored vaccine AZ1222 (AstraZeneca, vaccination only n = 19, and hybrid immunity n = 3). Blood was collected at the peak immunogenicity time point defined for each vaccine, for medians of eight, eight, 34, and seven days from the final dose, respectively. The demographic characteristic of the four groups of vaccinees is provided in Supplemental Table S1. The median age of the different cohorts was between 35-60 years old, with ages ranging from 21 to 77 years. For the vaccines including two doses within the primary immunization series (Pfizer, Moderna, and AstraZeneca), the median time between doses was three or four weeks for Pfizer and Moderna, respectively, and 66 days for the AstraZeneca vaccine, reflecting the policy in the United Kingdom where the AstraZeneca vaccinees resided.

### *mRNA and vector* SARS-CoV-2 *vaccines trigger functionally divergent antibody profiles*

Despite the fact that mRNA and vector SARS-CoV-2 vaccines each induce antibody titers and neutralizing antibodies (5, 11, 34, 35), significant differences have been noted between these vaccine platforms (36), in part attributable to divergent antibody levels. Despite the greater than 90% mRNA vaccine efficacy reported in early trials, real-world effectiveness declined to 40% and 75% for the Pfizer/BNT16b2 and Moderna mRNA1273 as VOCs spread globally (37, 38). Moreover, vectored vaccines conferred 65% and 75% efficacy in early trials with the Ad26.COV2.S and AZD1222 vaccines (5, 6), respectively, raise the possibility that differences beyond the overall levels of antibodies could explain efficacy differences among the vaccines. Thus, we performed comprehensive antibody profiling across four groups of vaccinees against the original D614G Spike antigen (wild-type, WT) in naïve individuals at the peak immunogenicity (**Figure 1A**). Antibody profiles were interrogated against the full spike, as well as the S1 domain-, S2 domain-, receptor-binding domain- (RBD), and N-terminal domain- (NTD). Striking differences were observed in the vaccine-induced antibody profiles across the four vaccine platforms (**Figure 1B)**, marked by lower overall titers and functionality in the vector vaccine platforms, robust functionality in the mRNA1273 immunized individuals, and high titers and antibody functions observed in the BNT162b2 vaccinees. Univariate comparison of spike-specific antibody levels across the vaccine platforms highlighted the expected elevated levels of IgG1, IgG2, IgG3, IgA, and IgM responses triggered by the mRNA vaccines compared to the vector vaccines (**Figure 1C**). Additionally, while BNT162b2 exhibited the highest overall Spike-specific IgG levels, more elevated IgA and IgM responses were observed in mRNA-1273 vaccinees than BNT162b2 recipients. Similarly, Ad26.COV2.S vaccine recipients exhibited higher IgA and IgM compared to individuals who received the entire course of the AZD1222 vaccine. These data point to both mRNA/vector and within platform differences in antibody profiles.

**Figure 1.**
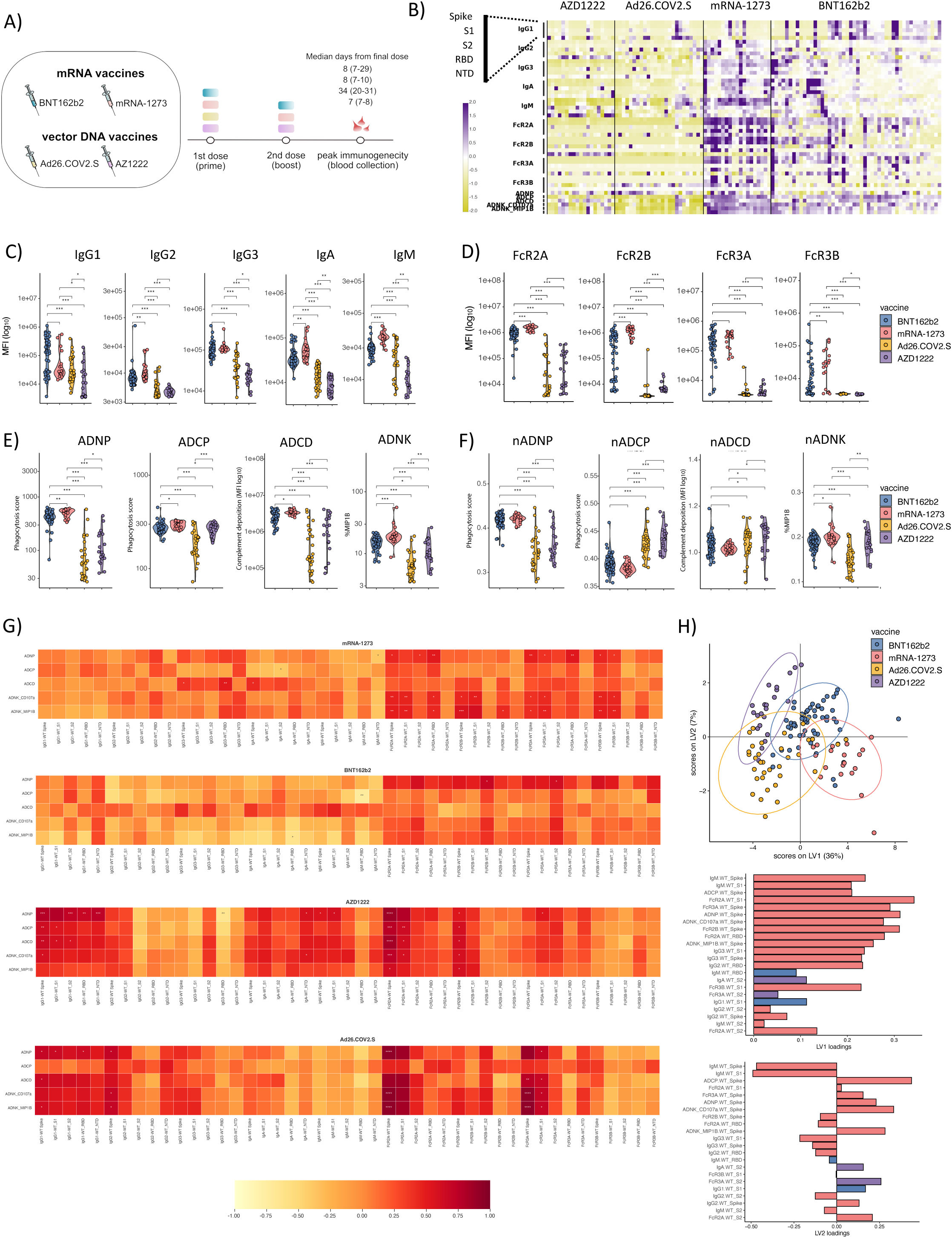
mRNA and vector DNA SARS-CoV-2 vaccines trigger functionally divergent antibody profiles. (**A)** Schematic representation of the study groups: # individuals received two doses on mRNA vaccines BNT162b2 (n = 48, in blue) or mRNA-1273 (n = 19, in pink), one dose of vector DNA Ad26.COV2.S vaccine (n = 25, in yellow) or two doses of vector DNA vaccine AZ1222 (n = 19, in violet). Blood was collected at the peak immunogenicity time point (with a median of 8, 8, 34, and 7 days from the final dose, respectively). **(B)** The heatmap summarizes the SARS-COV-2 WT Spike, S1, S2, RBD, and NTD-specific IgG1, IgG2, IgG3, IgA1, and IgM titers, as well as the ability of the SARS-CoV-2 specific antibodies to bind to the low-affinity Fcγ-receptors (FcγR2A, Fcγ2B, Fcγ3a, and Fcγ3b) across the individuals that received mRNA (BNT162b2 or mRNA-1273) or vector DNA (Ad26COV2.S or AZD122) vaccine at peak immunogenicity. Each column represents a different individual. Each row represents a distinct feature that was analyzed. **(C-F)** The violin plots show the univariate comparisons of WT Spike-specific **(C)** IgG1, IgG2, IgG3, IgA and IgM, **(D)** FcγR2A-, FcγR2B-, FcγR3A- and FcγR3B-binding levels and **(E)** functional properties, such as antibody-dependent neutrophil phagocytosis, cellular phagocytosis, complement deposition and NK cells activation across recipients of four different COVID-19 vaccine platforms. **(F)** The antibody-mediated effector functions were normalized to total IgG antibody level to highlight the effector properties of antibodies triggered by different CIVID-19 vaccines independent of antibody level. Each dot represents a single individual. Differences were defined using a Mann-Whitney test, and all p values were corrected for multiple comparisons using the Benjamin-Hochberg (BH) method, with *p* < 0.001 ***, *p* < 0.01 **, *p* < 0.05 *. **(G)** The correlation heatmap shows Spearman correlation matrices of the relationships between the antibody-mediated effector functions and SARS-CoV-2 WT Spike, S1, S2, RBD, and NTD antibody level and FcR binding properties at peak immunogenicity across four different COVID-19 vaccine platforms. Negative correlations are indicated in yellow-orange, and positive correlations are shown in red. **(H)** The partial least square discriminant analysis (PLS-DA) was used to visualize the separation between the COVID-19 vaccine platform based on the LASSO selected features. Each dot represents an individual vaccinee within the group. The bar graph shows the ranking of the LASSO selected features based on a Variable of Importance (VIP) score for LV1 and LV2 loadings.

To begin to capture other functional differences in the vaccine-induced humoral profiles, we next compared the Fcγ receptor (FcγR) binding profiles of SARS-CoV-2 specific antibodies at peak immunogenicity for each group. Robust FcγR-binding antibodies were observed for mRNA vaccinees, marked by significantly higher levels of Fcγ2A, Fcγ2B, and Fcγ3B binding in mRNA1273 vaccinees compared to BNT162b2 recipients. Moreover, vector-induced Spike-specific antibodies both bound to the activating opsonophagocytic Fcγ2A receptor, but both neither induce appreciable levels of Fcγ3A or Fcγ3B binding antibodies. Conversely, AZD1222 recipients elicited slightly higher levels of inhibitory-Fcγ2B binding antibodies compared to Ad26.COV2.S recipients (**Figure 1D)**, pointing to important differences across mRNA/vector platforms as well as within platforms in FcR binding profiles.

To determine whether the differences in FcR binding translated to functional differences, we next compared several Fc-mediated effector activities of vaccine-induced antibodies (**Figure 1E)**. As expected, mRNA vaccine induced more robust spike-specific antibody-mediated neutrophil phagocytosis (ADNP), cellular phagocytosis (ADCP), complement deposition (ADCD), and antibody-dependent Natural Killer (NK) activation (ADNK) activity. Within the mRNA vaccines, mRNA-1273 vaccinees exhibited the highest ADNP and ADNK activity. Additionally, between the vectored vaccines, AZD1222-triggered superior ADCP and ADNK activity compared to Ad26.COV2.S vaccination, highlighting differences between and within platforms.

However, given the striking differences in titers (**Figure 1C**) and FcR (**Figure 1D**) binding levels and the less pronounced differences in antibody effector functions across the platforms (**Figure 1E**), we next aimed to define the relationship of antibody levels to antibody effector function. Specifically, Spike-specific antibody functions were normalized to Spike-specific IgG levels to capture the functionality per antibody quality induced by each vaccine platform. Strikingly, normalization of ADNP, ADCP, ADCD, and ADNK to a total Spike-specific IgG level (**Figure 1F**) revealed the preferential induction of more ADNP and ADNK inducing antibodies by the mRNA platforms. Conversely, ADCP and ADCD were induced more effectively, on a per antibody level, by the vectored vaccines. Thus, on a per antibody level, distinct vaccine platforms may provide precise instructions to the evolving B cell immune response to elicit specific functional activities, that may be differentially controlled at the level of different isotype/subclass selection or Fc-glycosylation.

We next aimed to define the epitope-specific functional response across the vaccine platform. Specifically, the relationships between Spike-specific antibody functions and SARS-CoV-2 WT Spike-, S1-, S2-, RBD- and NTD-specific antibody levels and FcγR-binding were investigated (**Figures 1G**). Interestingly, IgG1 levels were strongly associated with antibody functions in mRNA1273 and Ad26.COV2.S immunized individuals. Conversely, FcR binding levels were tightly associated with antibody effector function in the BNT162b2 vaccinees, in addition to the mRNA1273 and Ad26.COV2.S vaccinees. Conversely, a more diffuse response was observed in AZD1222 vaccinees, with an interesting, stronger immunodominant relationship of S2-specific FcR binding associated with several antibody functions. Finally, IgA and IgM responses were solely associated with antibody effector functions in Ad26.COV2.S immunized individuals, pointing to striking differences in the overall architecture of the humoral immune response among the four vaccines.

Thus, to gain an overall appreciation for whether the vaccine profiles differ from one another, we finally used to integrate all the humoral data for each vaccine group and used Least Absolute Shrinkage and Selection Operator (LASSO) to conservatively reduce the overall features to the minimal number of vaccine measurements that could discriminate between all four COVID-19 vaccine-induced profiles at peak immunogenicity (**Figure 1H**). The data were then visualized using a Partial Least Squares Discriminant Analysis (PLS-DA). Latent variable 1 (LV1) separated vectored (left) and mRNA vaccines (right) from one another. Conversely, LV2 split the vaccines within the platforms, with BNT162b2 and AZD1222 segregating together (top) and mRNA1273 and Ad26.COV2.S segregating together (bottom). The latent space loading graphs bar graphs illustrated the minimal features that drove the separation between the vaccine profiles, marked by enhanced Spike- and S1-specific IgM, Fcγ2A, and Fcγ2B binding antibodies and effector functions such as ADCP, ADNP, and ADNK in mRNA vaccinated individuals. Conversely, RBD-specific IgM and S1-specific IgG1 were enriched in the vectored vaccines. Separation across the platforms was more nuanced, marked by enhanced ADCP, FcγR3a, ADNP, NK, and S2-specific FcγR2a binding in BNT162b2 and AZD1222, but more Spike specific IgM, FcγR2b, RBD-specific, IgG3, and S2-specific Ig FcγR2A in mRNA1273 and Ad26.COV2.S vaccinees. Thus, mRNA vaccination was marked by an overall magnitude increase in the functional quality of the humoral immune response, but differences in the overall functionality and epitope-specificity appeared to drive within platform differences in antibody profiles, which may help explain differences in real-world efficacy differences in the setting of emerging VOCs.

### Cross-vaccine antibody functional differences across VOCs

Despite the emergence of several VOCs that led to enhanced transmission due to neutralization escape, little is known about the functional quality of vaccine-induced antibodies with respect to VOCs. Thus, we next profiled the per-antibody vaccine-induced functional profile across the Alpha (B.1.1.7) and Delta (B.1.617.2) VOC Spikes (**Figure 2**). Similar to the profiles observed for the D614G Spike, on a per antibody level, mRNA vaccines induced higher ADNP and ADNK activity, albeit mRNA1273 induced slightly higher ADNK activating antibodies compared to BNT162b2 (**Figure 2A**). Conversely, ADCP and ADCD were higher on a per antibody level in the vectored vaccines than those in the mRNA vaccines, with AZD1222 inducing more elevated levels of ADCP per antibody level compared to Ad26.COV2.S vaccination.

**Figure 2.**
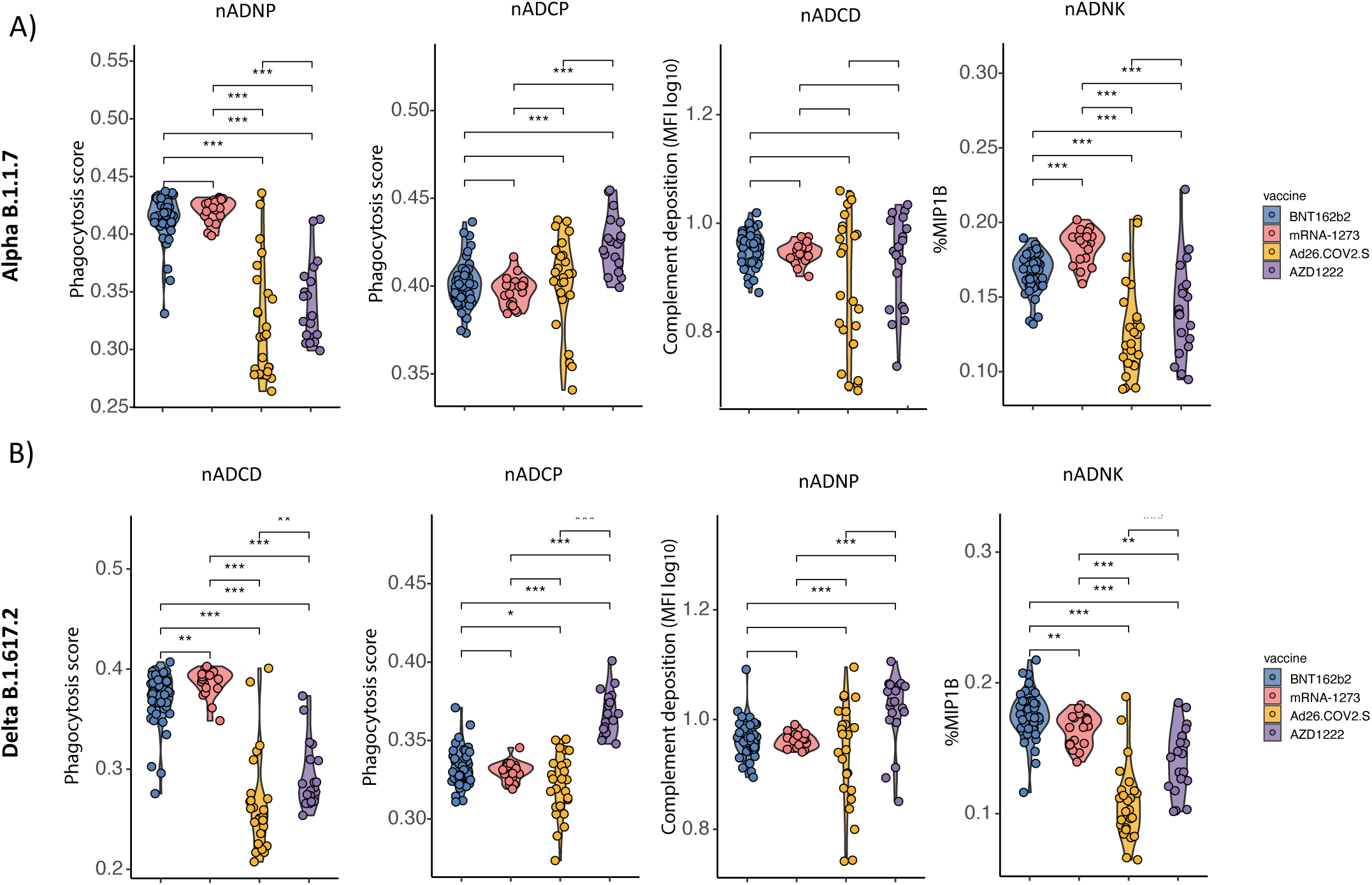
mRNA and vector DNA SARS-CoV-2 vaccines induce cross-reactive functional antibody across VOCs. Violin plots show the univariate comparisons of antibody-dependent neutrophil phagocytosis, cellular phagocytosis, complement deposition, and NK cells activation of **(A)** B.1.1.7 and **(B)** B.1.617.2 SARS-CoV-2 VOC Spike-specific antibodies normalized by the total IgG antibody level across four different COVID-19 vaccine platforms. Each dot represents a single individual. Differences were defined using a Mann-Whitney test, and all p values were corrected for multiple comparisons using the Benjamini-Hochberg (BH) method, with *p* < 0.001 ***, *p* < 0.01 **, *p* < 0.05 *.

Interestingly, the profiles of the Delta variant were distinct (**Figure 2B**), marked by higher levels of ADNP induced by mRNA1273 but higher levels of ADNK driven by BTN126b2, although both ADNP and ADNK were higher in mRNA vaccinees. For ADCP and ADCD, the profile was slightly different, with only AZD1222 inducing the highest levels of these opsonophagocytic functions compared to the other three platforms, pointing to substantial differences in functional humoral vaccine performance across VOCs.

### Characteristic of hybrid immunity and vaccination only cohort

Vaccination after natural SARS-CoV-2 infection, known as “hybrid immunity”, has been shown to increase the potency and breadth of humoral responses to SARS-CoV-2 (24, 25) and lead to enhanced real-world efficacy against VOCs (22, 23). However, it is unclear whether hybrid immunity can alter the functional quality of the humoral immune response. Thus, we next compared the cohort of individuals without prior SARS-CoV-2 infection (further described as “vaccination only”) to those with prior SARS-CoV-2 natural infection (hence “hybrid immunity”) who received (1) mRNA vaccines, including both the BNT162b2 (vaccination only n = 35, D1 n =35, D2 = 48; and hybrid immunity n = 9, D1 =9, D2 = 9) and the mRNA-1273 (vaccination only n = 19, D1 =19, D2 = 19; and hybrid immunity D1 n =1, D2 n = 1,); or (2) received either of the vectored vaccines, including the Ad26.COV2.S after 1^st^ dose (vaccination only n = 25, and hybrid immunity n = 8) and AZ1222 after (vaccination only n = 19, D1 = 30, D2 = 19; and hybrid immunity n = 3, D1 = 9, D2 = 3) (**Figure 3A**). Profiles were assessed after the first and second dose of the vaccines.

**Figure 3.**
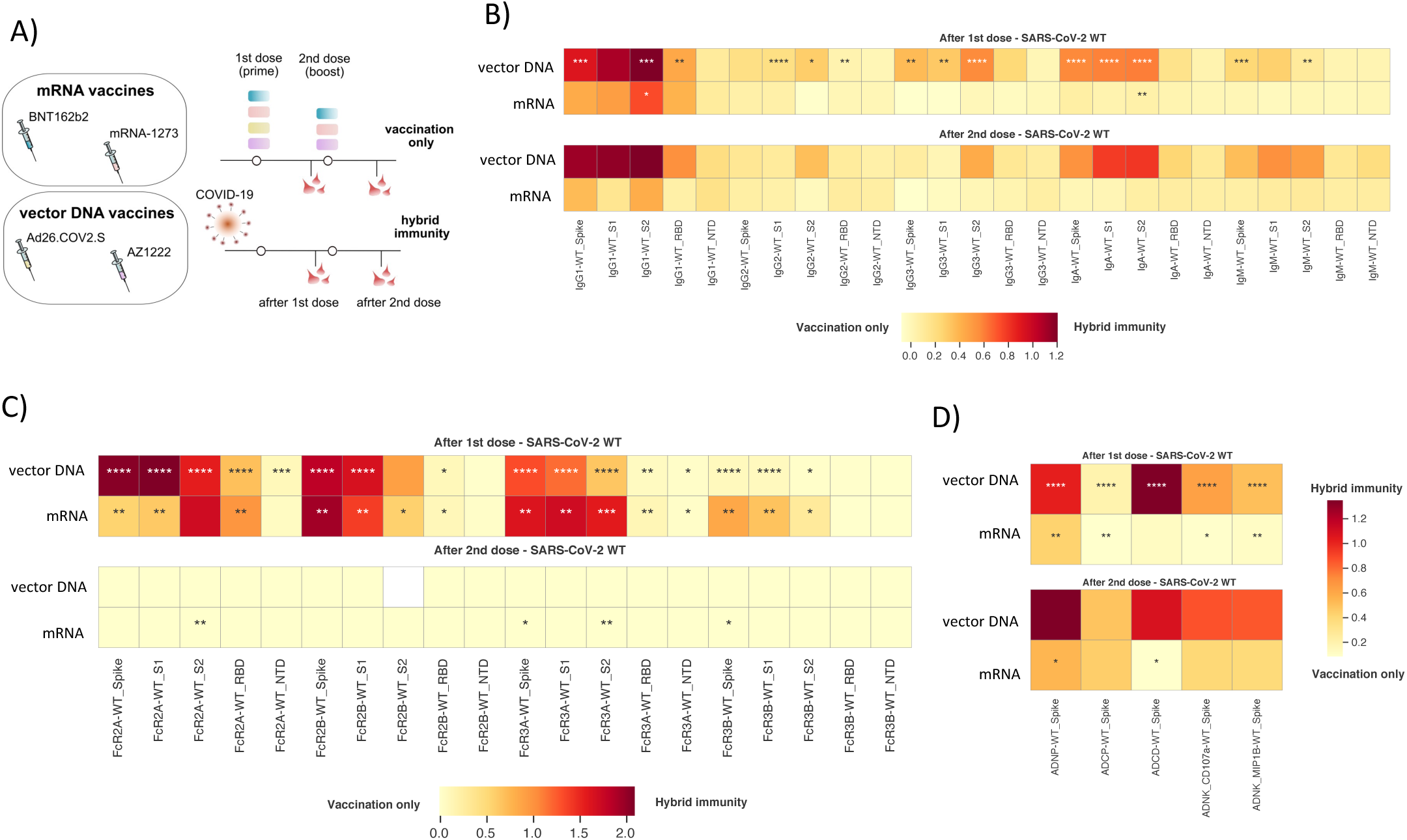
SARS-CoV-2 infection prior to one or two doses of mRNA and vector DNA vaccine enriches functional S2-specific antibody profile compared to vaccination only. **(A)** Schematic representation of the study groups: number individuals without (n = 98) and with prior COVID-19 (n = 21) received two doses on mRNA vaccines BNT162b2 (vaccination only n = 35, and hybrid immunity n = 9, in blue) or mRNA-1273 (vaccination only n = 19, and hybrid immunity n = 1, in pink), one dose of vector DNA Ad26.COV2.S vaccine (vaccination only n = 25, and hybrid immunity n = 8, in yellow) or two doses of vector DNA vaccine AZ1222 (vaccination only n = 19, and hybrid immunity n = 3, in violet). Blood was collected both after the first and second dose of a vaccine. **(B - D)** Heatmaps show the median delta value (Δ) in log10 scale between vaccination only and hybrid immunity for SARS-CoV-2 Spike-, S1-, S2-, RBD- and NTD-specific **(B)** IgG1, IgG2, IgG3, IgA, and IgM level **(C)** FcγR2A-, FcγR2B-, FcγR3A- and FcγR3B-binding levels, as well as **(D)** SARS-CoV-2 WT Spike-specific ADNP, ADCP, ADCD and ADNK after the first and second dose of four different COVID-19 vaccines. Differences were defined using a Mann-Whitney test, and all p values were corrected for multiple comparisons using the Benjamini-Hochberg (BH) method, with *p* < 0.001 ***, *p* < 0.01 **, *p* < 0.05 *.

Heatmaps were constructed using the SARS-CoV-2-specific antibody profiles depicting the median delta differences between individuals who only received the vaccine or who had hybrid immunity across vector and mRNA vaccines, with dark red indicating larger differences between groups (**Figure 3B-D)**. As expected, we observed enhanced augmentation of immunity after the 1^st^ dose in individuals who received a vectored vaccine (**Figure 3B**). Moreover, among the features, only two features in the mRNA vaccine-induced immune response were statistically significantly augmented in individuals with hybrid immunity, related to S2-specific IgG1, IgG3, and IgA levels. Interestingly, several antibody levels increased again in the vectored vaccinees after the second immunization, with a preferential, albeit not statistically significant, enrichment in S1 and S2-IgG1 and IgA responses. A trend towards a non-significant enrichment of Spike/S2 specific IgG1 was again observed in mRNA vaccinees with hybrid immunity. These data pointed to a more dramatic impact of hybrid immunity among vectored vaccinees, gaining both IgG1, IgG3, and IgA immunity and a selective increase in S2-specific immunity with mRNA vaccination.

To further define the functional impact of hybrid immunity, we next examined the differences in vaccine-induced FcR binding antibodies in individuals with hybrid immunity compared to the naïve population. Surprisingly, distinct from antibody isotype/subclass level changes, statistically significant augmentations were observed in FcR binding profiles across individuals who received a vector or mRNA vaccine (**Figure 3C**). Specifically, after the 1^st^ dose, individuals that received a vectored vaccine experienced a strikingly significant rise in Spike, S1, and S2-specific FcγR2a, FcγR2b, FcγR3a, and a significantly albeit more limited augmentation in FcγR3b binding antibodies. Interestingly, mRNA vaccinees also experienced a significant augmentation in Spike, S1, S2, and RBD-specific FcγR2a, FcγR2b, FcγR3a, and FcγR3b binding after the 1^st^ dose. Additional statistically significant although smaller improvements were noted in NTD-specific FcR binding in vectored volunteers. However, despite this remarkable improvement in functionality, no differences were noted in FcR binding after the 2^nd^ dose of vaccination in the hybrid immune group across the different vaccine platforms, suggesting that hybrid immunity may only afford an advantage in FcR binding after the 1^st^ dose of immunization.

However, to finally define whether hybrid immunity may modulate antibody effector function, we finally compared differences in antibody function after the 1^st^ and 2^nd^ dose of the vaccines (**Figure 3D**). After the 1^st^ does, a substantial enhancement was observed in all functions in vector immunized individuals, with the strongest augmentation in Spike-specific ADCD activity. Spike-specific ADNP, ADCP, and ADNK were also significantly increased in mRNA vaccinees after a single dose of the vaccine. Interestingly, after the 2^nd^ dose, individuals that received a vector still experienced a trend towards a large augmentation in ADNP, ADCD, and ADNK activity, suggesting that even after a 2^nd^ dose, individuals with hybrid immunity may harbor more robust antibody functionality to SARS-CoV-2 compared to individuals that received vectored vaccines alone. Interestingly, despite the fact the fold increase was smaller, hybrid immunity also showed increased levels of ADNP and ADCD among mRNA vaccinated individuals after the 2^nd^ dose, again pointing to potentially enhanced opsonophagocytic protection that may contribute to the improved real-world effectiveness observed in the setting of the previous infection.

To finally gain a more profound sense of the specific unique features of the humoral immune response induced by hybrid immunity, we performed a multivariate LASSO/PLSDA analysis (**Figure 4A-B**). Near-complete separation was observed between individuals with hybrid immunity compared to those solely vaccinated with either an mRNA (**Figure 4A**) or a vector vaccine after the firs dose. Interestingly, only three of a total of 40 features analyzed for each sample were sufficient to separate mRNA vaccinees with hybrid immunity or not. These features included a selective enrichment of S2-specific antibodies binding to FcγR2a, FcγR3b binding Spike-specific antibodies and S2-specific IgA in hybrid immunity. Conversely, hybrid immunity led to a highly statistically significant enrichment of multiple FcγR, IgA, and IgG1/2/3 responses in vector DNA vaccinated individuals, with a notable increase in S2-specific FcγR2a, IgG1, and IgA. After the 2^nd^ dose, mRNA vaccinee profiles continued to exhibit unique profiles in individuals with hybrid immunity, marked again by a selective augmentation of S2-specific FcγR2a and Spike-specific ADNP in hybrid immunity, compared to higher S1-specific IgG3 and Spike-specific ADCD among vaccinated only individuals (**Figure 4B**). Furthermore, paired nested mixed linear models were used to rank the features that differed most across hybrid and naïve immune profiles (**Figure 4 C-D**), highlighting the presence of S2-specific FcR binding antibodies as top predictors of hybrid immunity following a single (**Figure 4C**) or double dose (**Figure 4D**) of the mRNA vaccination, as well as among the top enriched features in hybrid immunity following double dose of vector vaccination. Hybrid immunity did augment the overall Spike-specific response following vector vaccination. Thus, these data argue that hybrid immunity extends the overall Spike-specific response following vectored immunity but selectively augments S2-specific immune responses following mRNA vaccination, potentially pointing to a breach of vaccine induced immunodominance that shifts the response to highly conserved regions of the Spike that may confer additional non-neutralizing protection against VOCs.

**Figure 4.**
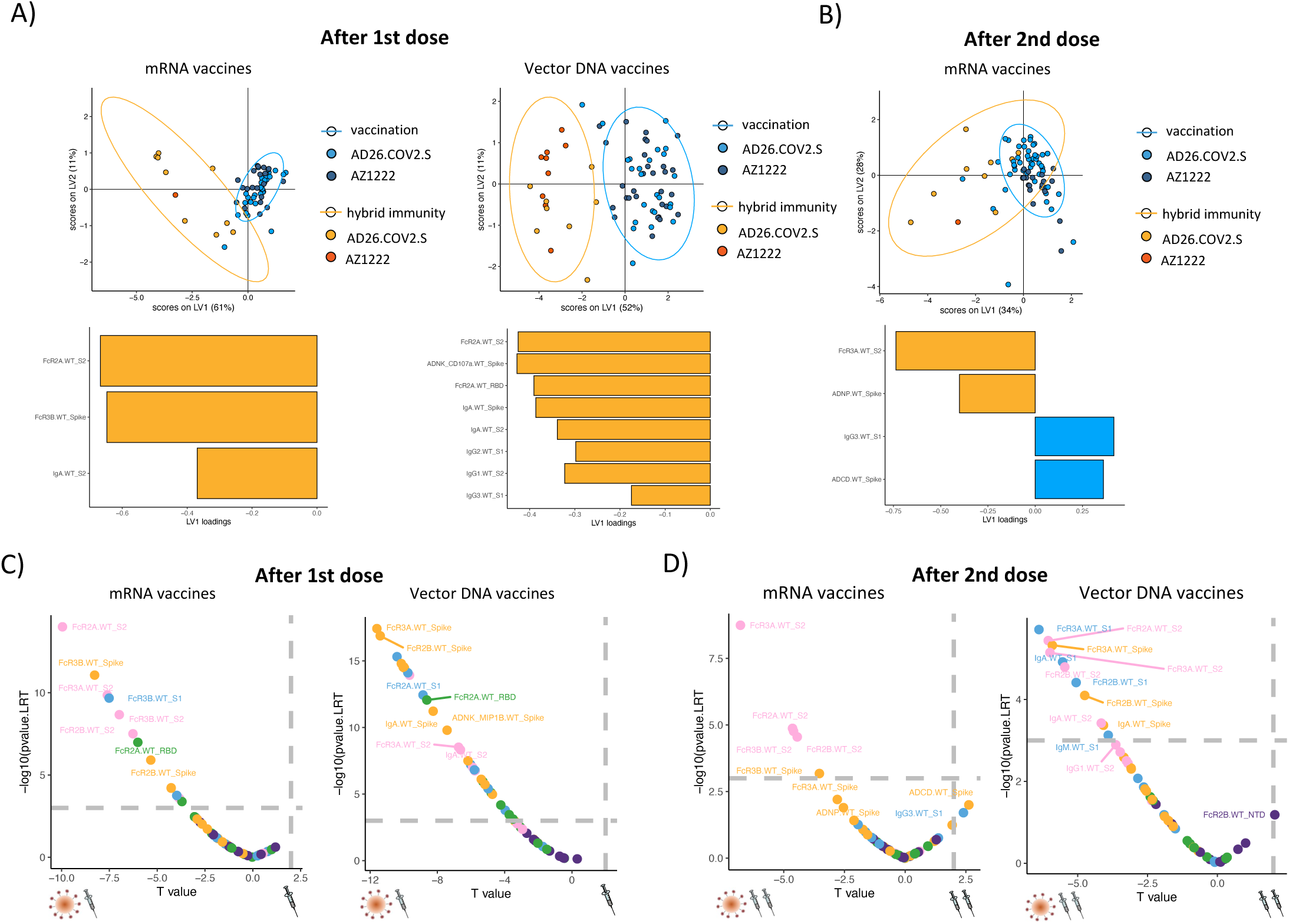
SARS-CoV-2 infection prior to one or two doses of mRNA and vector DNA vaccine induce specific unique features of the humoral immune response. The partial least square discriminant analysis (PLS-DA) was used to visualize the separation between vaccination only and hybrid immunity for mRNA and vector DNA vaccinees after **(A)** first and **(B)** second dose based on the WT SARS-Cov-2 LASSO selected features. Blue dots represent vaccination only for either BNT162b2 or AD26.COV2.S (light blue) and mRNA-1273 or AZ1222 (dark blue); yellow dots represent hybrid immunity for either BNT162b2 or AD26.COV2.S (light yellow) and mRNA-1273 or AZ1222 (orange). Each dot represents an individual vaccinee within the group. The bar graph shows the ranking of the LASSO selected features based on a Variable of Importance (VIP) score for LV1 and LV2 loadings, blue for vaccination only, and yellow for hydride immunity. Volcano plots of pairwise comparisons across vaccination only and hybrid immunity for vector DNA and mRNA vaccines after the first **(C)** and second dose **(D)** were controlled for age and gender of individuals. The x-axis represents the *t* value of the full model, and the *y* axis denotes the *P* values by likelihood ratio test comparing the null model and full model. The null/full model represents the association between each individual measurement (response) and all collected demographic information (see Methods). The horizontal gray dashed line denotes the *P-value* equals 0.05, and the vertical gray dashed line denotes a manually selected threshold (*t* values = 2).

### SARS-CoV-2 infection prior to one or two doses of mRNA and vector COVID-19 vaccine improves the functional breadth of SARS-CoV-2 VOC responses

Emerging data suggest that hybrid immunity may also expand the breadth of immunity to additional VOCs (24, 26-28). Given the unique expansion of S2-response observed with hybrid immunity, we next aimed to profile the overall landscape of antibody levels (**Figure 5A**), FcR binding profiles (**Figure 5B**), and functions (Figure 5C) across the mRNA and vectored vaccines. Focusing on Beta and Delta VOCs, we observed a statistically significant expansion of both Beta and Delta Spike-specific responses in hybrid vector-immunized individuals, with an augmentation, albeit non-significant in the Beta-RBD, but a lesser but significant augmentation in Delta-RBD binding IgG1 levels, suggesting that the majority of hybrid induced antibody responses may be directed outside of the RBD. Interestingly, hybrid immunity after the 1^st^ dose of the vector was also accompanied by an increase in IgG1, IgA, and IgM responses preferentially, pointing to a broader enhancement of immunity. This augmentation was less significant in hybrid-immune individuals after the 1^st^ dose of mRNA vaccination, although low, less significant increases were noted in IgG1 responses to the Beta and Delta Spikes and RBDs. Moreover, after the 2^nd^ dose of the vaccine, an increase in binding antibodies was noted in hybrid-immune individuals after vector vaccination. Still, limited increases in binding levels were noted in the 2^nd^ mRNA vaccinated hybrid immune individuals, except for a slight significant increase in D614G and Beta-Spike, not RBD, IgG3 responses.

**Figure 5.**
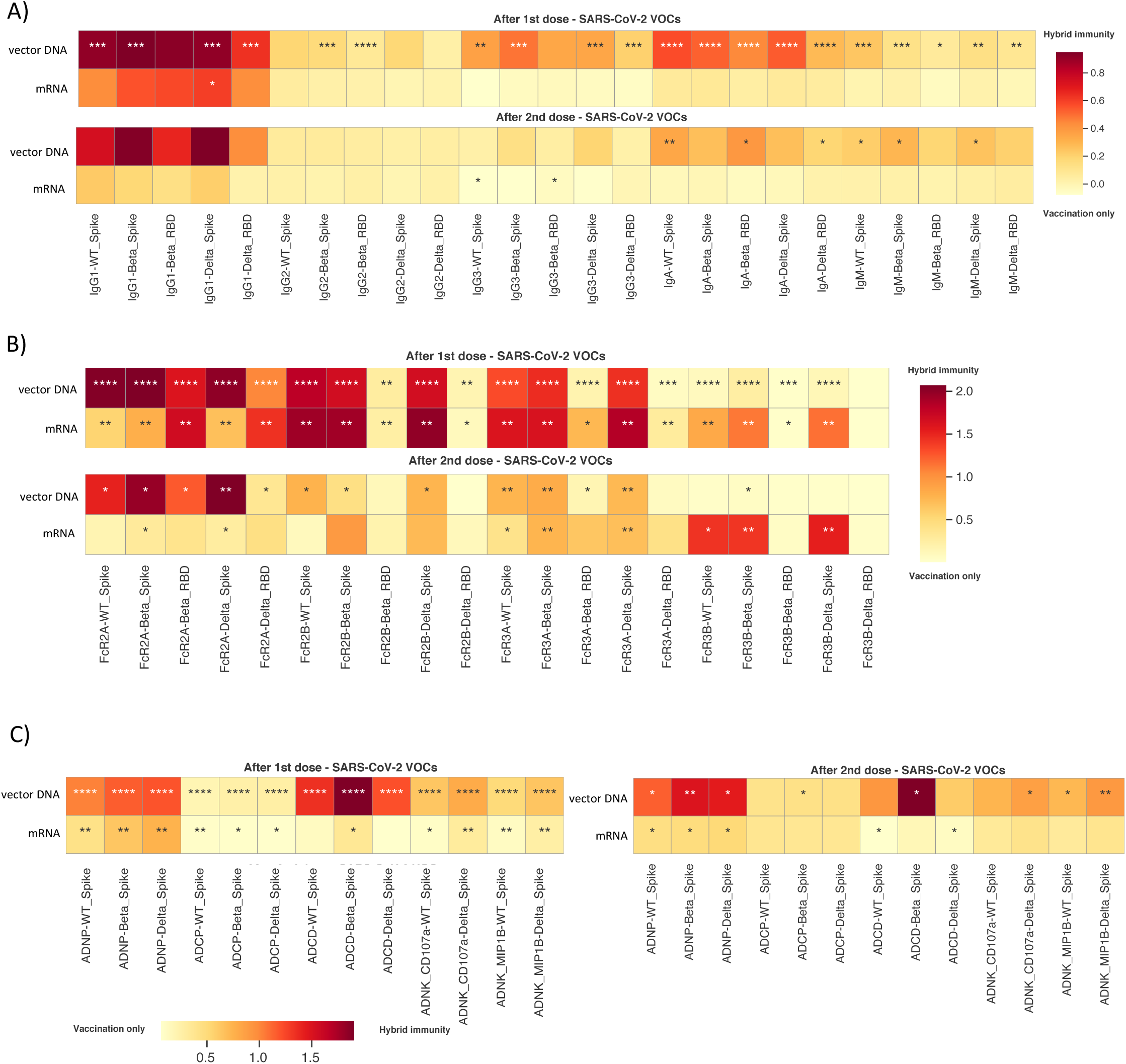
SARS-CoV-2 infection prior to one or two doses of mRNA and vector DNA vaccine improves the functional breadth of SARS-CoV-2 VOCs antibody response. **(A - C)** Heatmaps show the median delta value (Δ) in log10 scale between vaccination only and hybrid immunity for SARS-CoV-2 WT, B.1.1.7, B.1.351, P1, and B.1.617.2 Spike- and RBD - specific **(A)** IgG1, IgG2, IgG3, IgA and IgM level, **(B)** FcγR2A-, FcγR2B-, FcγR3A- and FcγR3B-binding levels, as well as **(C)** SARS-CoV-2 WT Spike-specific ADNP, ADCP, ADCD and ADNK after the first and second dose of four different COVID-19 vaccines. Differences were defined using a Mann-Whitney test, and all p values were corrected for multiple comparisons using the Benjamini-Hochberg (BH) method, with *p* < 0.001 ***, *p* < 0.01 **, *p* < 0.05 *.

In contrast to the more limited increases in antibody/isotype titers, a highly statistically significant increase in FcR binding antibodies was noted across both platforms after the 1^st^ immunization in hybrid-immune individuals, marked by higher levels of FcR2A and FcR3b binding antibodies to all VOC antigens, selectively to whole Spike VOC antigens for FcR2b and FcR3a. Similar although less potent augmentation was observed in hybrid-immune individuals after the 1^st^ dose of mRNA, highlighting the preferential increase in FcR binding antibodies to whole spike VOC antigens. Similarly, functional profiling across VOCs pointed to an overall significant expansion of all antibody functions following a single vector vaccination (**Figure 5C**), most dramatically observed for ADCD and ADNP. However, a single dose of mRNA vaccination also drove enhanced functionality, particularly ADNP activity, in individuals that were previously infected. Even a second dose of vectored vaccination further enhanced ADNP and ADCD to the Beta-Spike. Conversely, more limited functional augmentation was observed with the 2^nd^ dose of mRNA vaccination, albeit the augmentation was significant for ADNP and ADCD. Thus, these data suggest that hybrid immune augmentation of particular antibody effector functions across VOCs may contribute to enhanced control and clearance of the infection and ultimately in persistent attenuation of disease in the setting of neutralization escape.

In summary, we have highlighted herein previously unappreciated Fc-profile differences across mRNA vaccine as well as adenoviral vector vaccines, in addition to recapitulating previously observed differences in neutralizing antibody levels and binding antibody titers between vaccine platforms. Despite the lower antibody titers induced by vectored-vaccines similar functional opsonophagocytic functions were induced at a per antibody level, potentially conferring higher levels of protection against VOCs via non-neutralizing antibody mechanisms. Moreover, while the platforms benefited from hybrid immunity at differential levels, all platforms led to augmented S2-specific immunity that may play an essential role in VOC-mediated non-neutralizing control and attenuation of disease.

## Discussion

The COVID-19 pandemic led to a revolution in vaccine development, resulting in the large-scale testing of several novel vaccine platforms and adjuvants in response to this rapidly spreading and often unpredictable pathogen. This near head-to-head testing of novel vaccines highlighted differences in efficacy induced by distinct platforms (9, 39-41). However, as the pandemic evolved, waning immunity and the evolution of neutralization resistant variants of concern (VOC) challenged the assumed correlates of protection, as neutralizing antibody titers were no longer a robust predictor of protection against severe COVID-19. Instead, transmission increased globally in the absence of a concomitant rise in severe disease and death in vaccinated populations, suggesting that other aspects of the vaccine-induced immune response are likely to be key to protection against disease. Differences in neutralizing titers have not reliably predicted discrepancies in real-world effectiveness between mRNA vaccines (37, 38). Therefore, antibody levels and T cell immune responses, robustly induced by vectored vaccines, have not been directly predictive of differences in vaccine efficacy (42-44). Moreover, emerging real-world data pointed to the highest levels of efficacy against COVID-19 among individuals that had been previously infected with SARS-CoV2 prior to vaccination, also termed hybrid immunity (23, 43, 45). The identification of platform-specific immune programming differences and how these may be tuned by hybrid immunity offers a unique opportunity to define the immunologic correlates of protection to guide rational boosting and next generation vaccine design against newly emerging VOCs.

Several studies have shown an association between antibody binding titers and neutralization with protection across Phase3 SARS-CoV-2 vaccine trials (46, 47), as well as clinical efficacy against primary infection with SARS-CoV-2 after vaccination (16, 48), suggesting that post-immunization antibody levels can serve as a measure for short-term protection against symptomatic infection as well as hospitalization and death across diverse vaccine platforms (16, 49, 50). While it has been observed that vaccine-induced neutralizing antibodies decrease rapidly over time (51-55), binding antibodies appear to be more durable (17, 56), potentially pointing to a role for non-neutralizing antibody functions in long-term protection against disease. Additionally, in the setting of VOCs able to evade neutralizing antibodies (57-59), emerging data suggest that vaccine-induced antibodies continue to recognize VOC Spike antigens further arguing that additional vaccine-induced antibody mechanisms likely contribute to protection (60-62). Despite our growing appreciation for the role of innate immune recruiting Fc-effector antibody functions in the resolution of severe disease (20, 21), monoclonal therapeutic efficacy (19), and survival after convalescent plasma therapy (63), little is known about the differences in Fc-effector functions across vaccine platforms as well as how they may evolve in the setting of hybrid immunity.

As expected, mRNA vaccines induce higher antibody titers, Fc-receptor binding profiles, and antibody effector function than vectored vaccines. However, even among the mRNA vaccines differences were noted in the magnitude of the functional humoral immune response. Higher IgG1 as well as IgG3 and IgA responses induced by the BNT162b2 and mRNA1273, respectively, were linked to enhanced FcR binding, ADNP, ADCP, ADCD, and ADNK, marking significant differences in antibody responses across SARS-CoV-2 mRNA vaccines platforms, potentially due to diversity in mRNA doses, intervals between vaccine injections, and lipid-nanoparticle formulation. Even across the adenoviral platforms, Ad26.CoV2.S induced higher titers compared to AZD1222, despite the fact that Ad26.CoV2.S was only administered as a single dose. Yet, AZD1222 induced higher FcγR2b binding antibodies as well as higher ADNP, ADCP, and ADNK activating antibodies compared to Ad26.CoV2.S, pointing to differences in the ability of distinct adenoviral vectors to drive diverse functional antibody profiles. Yet, most surprising was the observation that normalization of antibody effector function by antibody titers pointed to higher quality functional antibodies induced by vectors vaccines compared to mRNA, with superior levels of ADCP and a trend towards higher ADCD induced by vectors compared to mRNA vaccines. However, the higher real-world efficacy of mRNA vaccines compare to vector DNA, might suggest the cumulative effect of binding and functional antibodies responsible for protection from SARS-CoV-2. Whether this is related to the internal pattern recognition signal or innate cytokine responses induced by vectors compared to mRNA vaccines remains unclear. However, it may indicate the potential importance of using vectors as a prime in possible future heterologous vaccine strategies. The unique antibody functional profile generated by each vaccine is likely associated with the distinct inflammatory cues induced at the time of immunization, pushing T cell help and B cell responses towards specific class switch recombination and Fc-glycosylation profiles that collectively shape the overall FcR binding and effector profiles of vaccine-induced polyclonal swarms of antibodies. Further dissection of these distinct signals may provide critical insights for future vaccine design efforts to elicit highly specialized antibody functional profiles to maximize disease protection.

Emerging real-world efficacy data suggests that vaccination following infection with SARS-CoV2 results in the greatest level of protection against severe COVID-19 disease (23, 43, 45). This hybrid immune response quantitatively and qualitatively improved B cell and T cell responses (64-66). Here we observed that hybrid immunity also shapes antibody effector functions, with dramatic improvements in antibody response and FcR binding after a single dose of vectored vaccines and a trend towards an additional augmentation of titers and function following the second dose of AZD1222. Hybrid immunity led to a significant increase in S2-specific IgG and IgA titers for mRNA vaccines, pointing to a selective expansion of immunity to the conserved portion of the Spike antigen, with only trends in IgG1 titer and increases after the second dose of mRNA. Conversely, ADNP increased significantly after a first and second dose of mRNA vaccines in individuals with hybrid immunity, a function that has been specific selectively enriched among survivors of severe COVID-19 and vaccinated macaques that resisted SARS-CoV-2 infection (67). Moreover, after adjusting for demographics, S2-specific FcR binding was selectively enriched among mRNA vaccinees with hybrid immunity but also observed following AZD1222 vaccination. This selective improvement of S2-specific immunity may point to two critical phenomena: (1) vaccination may promote largely immunodominant immunity to the S1 domain (including the RBD and NTD), whereas infection may help expand the response to the more conserved and less immunogenic S2 domain that is not as exposed mainly in the stabilized spike antigens included in the mRNA vaccines; and (2) the expansion of immunity to the conserved S2 domain may be key to promoting cross-VOC immunity due to the higher conservation of S2 compared to other domains of the Spike antigen (68-70). Increased S2-specific antibodies able to mediate Fc effector functions were previously observed among SARS-CoV-2 survivors (20, 71) and in pre-pandemic children (*29*) who are typically spared from SARS-CoV-2 (72, 73). Due to their lower neutralizing potency (74), S2-specific antibodies provide protection against disease in an Fc-dependent manner (71), consistent with the mechanism by which hybrid immunity likely offers enhanced protection against VOCs. Specifically, the expansion of S2-specific responses with enhanced FcR binding likely promotes rapid capture and clearance of the virus after transmission. Because innate immune cells able to drive antibody-mediated clearance and killing are found more abundantly in the lower respiratory tract, these antibodies are unlikely to promote restriction in the upper respiratory but contribute to disease attenuation in collaboration with T cell immunity. Thus, given the structural conservation of the S2-domain, as well as the neutralization and cross-reactive potential of S2 antibodies, might reduce the influence of sequence-altering mutations and therefore could improve vaccine efficacy against seasonal circulating common cold coronaviruses and newly emerging VOCs.

There are some limitations to our study. We did not have large numbers of hybrid immune subjects that received the mRNA-1273 and AZ1222 vaccines and thus were unable to look for the impact of hybrid immunity in tuning specific vaccine platforms. Instead, we combined hybrid vaccinees that received an mRNA vaccine or DNA vaccine, given that the four-way discriminant analysis clearly highlighted that the platforms contributed to the greatest amount of variation in antibody profiles (LV1). However, future investigation, based on individual platforms may reveal additional differences in hybrid immune effects given discrepancies in antigen stabilization or persistence following immunization. Additionally, this study was unable to address issues related to immune profiles over time. Yet despite our results show functionally divergent antibody profiles triggered by mRNA and adenoviral vectored vaccine platforms in naïve individuals, as well as the unique effect of hybrid immunity on expanding both mRNA and vector vaccine-induced immunity to the conserved S2-domain. These data highlight the unique programming effects across the vaccine platforms as well as the potentially critical importance of promoting functional humoral immunity to more conserved regions of the beta-coronaviruses, collectively pointing to new opportunities to develop next-generation COVID-19 vaccines or mix-and-match combinations able to drive the most effective immunity existing and newly emerging VOCs.

## Methods

### Cohort description

Samples were collected post-vaccination from groups of individuals receiving the vaccine as part of their government’s national rollout campaigns with the verbal consent of participants (17). Samples from Latvia and South Africa vaccinees were obtained as part of a previous study of HCWs in pediatric facilities originally initiated at Great Ormond Street Hospital (COSTARS, IRAS 282713, ClinicalTrials.gov Identifier: NCT04380896). Ethics approval was obtained locally by the lead investigators of each site. In the UK, volunteers who were part of the COSTARS Study, as well as others who had received vaccines as part of the government rollout altruistically, agreed to donate serum to help evaluate an assay for measuring post-vaccine immunity being run the UCL laboratory. Vaccinees received one of four vaccines depending on local availability. In Latvia and South Africa, serum was aliquoted, given a unique identifier, and stored frozen until batch shipping to the WHO International Reference laboratory for Pneumococcal Serology at University College London, London, UK. Local UK samples had serum extracted and were stored frozen until batch tested.

### Luminex profiling

Antibody isotyping and Fcγ-receptor (FcγR) binding were conducted by multiplexed Luminex assay, as previously described (75, 76). Briefly, SARS-CoV-2 antigens were used to profile specific humoral immune responses. Antigens were coupled to magnetic Luminex beads (Luminex Corp) by carbodiimide-NHS ester-coupling (Thermo Fisher). Antigen-coupled microspheres were washed and incubated with plasma samples at an appropriate sample dilution (1:500 for IgG1 and all low-affinity Fcγ-receptors, and 1:100 for all other readouts) for 2 hours at 37°C in 384-well plates (Greiner Bio-One). The high-affinity FcR was not tested due to its minimal role in tuning antibody effector function (77). Unbound antibodies were washed away, and antigen-bound antibodies were detected by using a PE-coupled detection antibody for each subclass and isotype (IgG1, IgG3, IgA1, and IgM; Southern Biotech), and Fcγ-receptors were fluorescently labeled with PE before addition to immune complexes (FcγR2a, FcγR3a; Duke Protein Production facility). After one hour of incubation, plates were washed, flow cytometry was performed with an IQue (Intellicyt), and analysis was performed on IntelliCyt ForeCyt (v8.1). PE median fluorescent intensity (MFI) is reported as a readout for antigen-specific antibody titers.

### Effector functional assays

Antibody-dependent cellular phagocytosis (ADCP), antibody-dependent neutrophil phagocytosis (ADNP), antibody-dependent complement deposition (ADCD), and antibody-dependent NK activation assays were performed as previously described (Bulter et al., 2019; Karsten et al., 2019; Fischinger et al., 2019, Chung et al., 2015).

SARS-CoV-2 Spike proteins were coupled to yellow-green (505/515) or red-orange (565/580) fluorescent Neutravidin-conjugated beads (Thermo Fisher) for ADCP/ADNP and ADCD, respectively. Immune complexes were formed by incubating the diluted pooled samples (ADCP and ADNP 1:100 dilution) with the antigen-coupled beads for two h at 37°C. For ADCP, 1.25×10^5^ THP-1 cells/mL were added to the immune complexes and incubated for approximately 18 h at 37°C. After the incubation, THP-1 cells were washed and fixed with 4% paraformaldehyde (PFA) (Alfa Aesar). For ADNP, the immune complexes were incubated with 5 ×10^5^ cells/ml of RBC-lysed whole blood for one h at 37°C. After incubation, cells were washed and stained for CD66b+ (Biolegend) to identify neutrophiles and then fixed in 4% PFA.

For ADCD, the antigen-coupled beads were incubated with the diluted pooled samples (1:10 dilution) for two h at 37°C to form immune complexes. The immune complexes were washed, and lyophilized guinea pig complement (Cedarlane) in gelatin veronal buffer with calcium and magnesium (GBV++) (Boston BioProducts) was added for 30 min (complement was reconstituted according to manufacturer’s instruction). The deposition of complement was detected by fluorescein-conjugated goat IgG fraction to guinea pig Complement C3 (Mpbio).

All the assays were acquired by flow cytometry with iQue (Intelluicyt), and the analysis was performed using IntelliCyt ForeCyt. The phagocytosis score was calculated (% cells positive × Median Fluorescent Intensity of positive cells) for ADCP and ADNP. ADCD was reported as the median of C3 deposition.

For antibody-dependent NK cell degranulation, SARS-CoV-2 antigens were coated to 96-well ELISA at the protein concentration of 2 ug/ml, incubated at 37°C for 2hrs and blocked with 5% BSA at 4°C overnight. NK cells were isolated from whole blood from healthy donors (by negative selection using RosetteSep (STEMCELL) then separated using a ficoll gradient. NK cells were rested overnight in media supplemented with IL-15. Serum samples were diluted at 1:25. After blocking, samples were added to coated plates, and immune complexes were formed for two hours at 37°C. After the two hours, NK cells were prepared (antiCD107a– phycoerythrin (PE) – Cy5 (BD, 1:40, clone: H4A3), brefeldin A (10 µg/ml) (Sigma), and GolgiStop (BD)), and added to each well. for 5 hours at 37°C. The cells were stained for surface markers using anti-CD56 PE-Cy7 (BD, 1:200, clone: B159), anti-CD16 APC-Cy5 (BD, 1:200, clone: 3G8), and anti-CD3 PacBlue (BD, 1:800, UCHT1) and permeabilized with FIX & PERM Cell Permeabilization Kit (Thermo Fisher). After permeabilization, cells were stained for intracellular markers MIP1β (BD, 1:50, clone: D21–1351) and IFNγ (BD, 1:17, clone: B27). The flow cytometry was performed. NK cells were defined as CD3-CD16+CD56+ and frequencies of degranulated (CD107a+), INFγ+ and MIP1β+ NK cells determined on an iQue analyzer (Intellicyt).

### Univariate comparison

Data analysis was performed using R version 4.1.2. Univariate comparisons between four different vaccine platforms were performed using the Wilcoxon-signed rank test followed by Benjamini-Hochberg (BH) correction. The visualization was performed by the function “ggplot” of R package “ggplot2” (3.3.5), and the *P-value* was estimated by the function “wilcox.test” and “p.adjust” in the R package “stats”(4.1.2) and labeled by the function “stat_pvalue_manual” in the R package “ggpubr” (0.4.0). The heatmap showing the difference between median of hybrid immunity group vs. that of vaccination only group in log10 scale was plotted by “heatmap” function of python module “seaborn” (0.11.2), the *P* value was estimated by the function “mannwhitneyu” of python module “scipy.stats” (1.8.0), followed by the Benjamini-Hochberg procedure of multiple testing correction using function “multipletests” of python module “statsmodels.stats.multitest” (0.13.2).

### Multivariate classification

Classification models were trained to discriminate between vaccination only and hybrid immunity individuals using all the measured antibody responses. Prior to analysis, all data were normalized using z-scoring. Models were built using a combination of the least absolute shrinkage and selection operator (LASSO) for feature selection and then classification using partial least square discriminant analysis (PLS-DA) with the LASSO-selected features (54) using R package “ropls” version 1.26.4 (Thévenot et al., 2015) and “glmnet” (4.1.3). Model accuracy was assessed using five-fold cross-validation. For each test fold, LASSO-based feature selection was performed on logistic regression using the training set for that fold. LASSO was repeated 10 times, features selected at least 7 times out of 10 were identified as selected features. PLS-DA classifier was applied to the training set using the selected features, and prediction accuracy was recorded. The first two latent variables (LVs) of the PLS-DA model were used to visualize the samples.

### Correlation analysis

Spearman correlations were used to correlation between antibody titers and functional responses and were performed using function “spearmanr” of python module “scipy.stats” (1.8.0). The significance of correlation was adjusted by the Benjamini-Hochberg procedure of multiple testing correction using function “multipletests” of python module “statsmodels.stats.multitest” (0.13.2).

### Defining signatures of previous infection while also controlling for potential cofounders

We accessed the significance of the association between measured antibody levels and the previous infection state by controlling for collected potential cofounders using two nested mixed linear models (null and full model) without/with demographical data, i.e., age and gender in this study. We fit two linear mixed models and estimated the improvement in model fit by likelihood ratio testing to assess how many measurements have a significantly better fit with the full model at a threshold of <0.05.

#### Null model

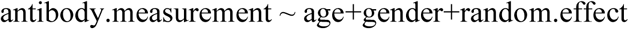

#### Full model

antibody.measurement ∼ age+gender+previous.infected+random.effect Likelihood ratio test LRT=-2*ln(MLE in Full model MLT in Null model)∼λ2, MLE denotes maximum likelihood estimation and MLT denotes maximum likelihood ratio.

Here, we consider the measurement of IgG1 titer in HA_control as the random effect. The R package “lme4” was used to fit the linear mixed model to each measurement and test for measurement across the contrast of interest. The *P* value from the likelihood ratio test and *t* value of previous infected state in full model were visualized as volcano plot using the “ggplot” function in R package “ggplot2”.

### Data availability

All anonymized data collected during the trial and associated with this study can be provided. Request should be directed to. Data requestors will need to sign a data access agreement to gain access, and access will be granted for non-commercial research purposes only.

## Acknowledgments

We thank Nancy Zimmerman, Mark and Lisa Schwartz, an anonymous donor (financial support), Terry and Susan Ragon, and the SAMANA Kay MGH Research Scholars award for their support. We acknowledge the support from the Ragon Institute of MGH, MIT, and Harvard (G.A) the Massachusetts Consortium on Pathogen Readiness (MassCPR) (G.A.), and the National Institutes of Health (3R37AI080289-11S1, R01AI146785, U19AI42790-01, U19AI135995-02, U19AI42790-01, 1U01CA260476 – 01, CIVIC75N93019C00052) (G.A.). H.Z. was funded by a UK NIHR GECO award (GEC111).

## Author Contribution statement

P.K., Y.X., D.A.L, D.G. and G.A. analyzed and interpreted the data. P.K. and J.S.L., performed Luminex and functional assays. Y.X. performed the analysis. H.Z. and D.Z. helped procure the cohorts. M.J. and D.G. was responsible for all aspects of the clinical study and sample collection. P.K. and G.A. drafted the manuscript. All authors critically reviewed the manuscript.

## Competing Interests Statement

G.A. is a founder and equity holder for Seromyx Systems Inc., an employee and equity holder for Leyden Labs, and has received financial support from Abbvie, BioNtech, GSK, Gilead, Merck, Moderna, Novartis, Pfizer, and Sanofi. G.A.’s interests were reviewed and are managed by Massachusetts General Hospital and Partners HealthCare in accordance with their conflict-of-interest policies. All other authors have declared that no conflict of interest exists.

## Notes

### Author Declarations

Samples from vaccinees in the UK, Latvia, and South Africa were obtained as part of a previous study of healthcare workers in pediatric facilities originally initiated at Great Ormond Street Hospital (COVID-19 Staff Testing of Antibody Responses Study (Co-STARS)), the Integrated Research Application System (IRAS) project ID: 282713, ClinicalTrials.gov Identifier: NCT04380896). Ethics approval was obtained locally by the lead investigators of each site. Ethical approvals were given by: UK: Health Research Agency in England and Health and Care Research Wales South Africa: Human Research Ethics Committee, Faculty of Health Sciences, University of Cape Town Latvia: Riga Stradinas Universitaes, Etikas Komiteja

